# SeGA-GNN: Semantically Gated Augmented Graph Neural Networks for Wearable-Based Emotion Detection

**DOI:** 10.64898/2026.05.29.26354434

**Authors:** Furkan Kurt, Seyma Nur Subasi, Ezgi S. Yakisan, Abdulhamit Subasi

**Affiliations:** Department of Information Science and Technology, University at Albany, SUNY, NY, USA; Büyükçekmece Mimar Sinan Devlet Hastanesi, Mimar Sinan Merkez Mah. D100 Karayolu, No:62 34535, Büyükçekmece, Istanbul, Turkiye; Department of Emergency Health Services, Republic of Turkiye Ministry of Health, Turkiye

**Keywords:** Mood Prediction, Emotion detection, Multimodal learning, Wearable sensing, Fitbit data, Digital phenotyping, Graph Neural Networks (GNN), Heterogeneous graphs, Large Language Models (LLM), Semantic embeddings, Multimodal data fusion, Mental health monitoring, Affective computing, Knowledge graphs, Time-series analysis, Explainable AI

## Abstract

**Background:** Wearable technologies enable scalable and continuous monitoring of emotional states through passive sensing of physiological and behavioral signals. However, conventional learning approaches often struggle to model the complex temporal, contextual, and relational dependencies underlying human emotions. To address these limitations, we propose a graph-based framework that represents multimodal wearable observations as heterogeneous knowledge graphs enriched with semantic information derived from Large Language Models (LLMs), enabling richer contextual understanding beyond raw sensor measurements.

**Methods:** We constructed a heterogeneous knowledge graph using multimodal Fitbit physiological signals and affective self-report data collected from 45 users. Framing mood prediction and emotion detection was formulated as both binary and ternary node classification tasks. We evaluated five baseline heterogeneous Graph Neural Network (GNN) architectures and compared them with the proposed Semantically Gated Augmented Graph Neural Network (SeGA-GNN) framework, which dynamically integrates LLM-generated semantic embeddings into graph representations through a gated cross-modal fusion mechanism.

**Results:** The baseline GNN models aachieved strong performance, with classification accuracies ranging from 0.7525 to 0.9739 for binary classification and 0.6249 to 0.9699 for ternary classification. The proposed SeGA framework consistently improved predictive performance across most architectures. In particular, semantic augmentation transformed the HAN model from moderate baseline performance into near-perfect emotion recognition capability, achieving SeGA-HAN Accuracy = 0.9988 and AUC = 1.0000 for binary classification and Accuracy = 0.9979 and AUC = 1.0000 for ternary classification.

**Discussion and Conclusion:** Integrating LLM-derived semantic contextualization into heterogeneous graph learning enables effective modeling of contextual information that is not directly captured by wearable physiological signals alone. The proposed SeGA-GNN framework demonstrates that adaptive semantic fusion substantially improves the accuracy, robustness, and interpretability of wearable-based emotion detection. These findings establish a promising direction for next-generation wearable affective computing systems and intelligent emotion-aware applications.

**Graphical Abstract:** 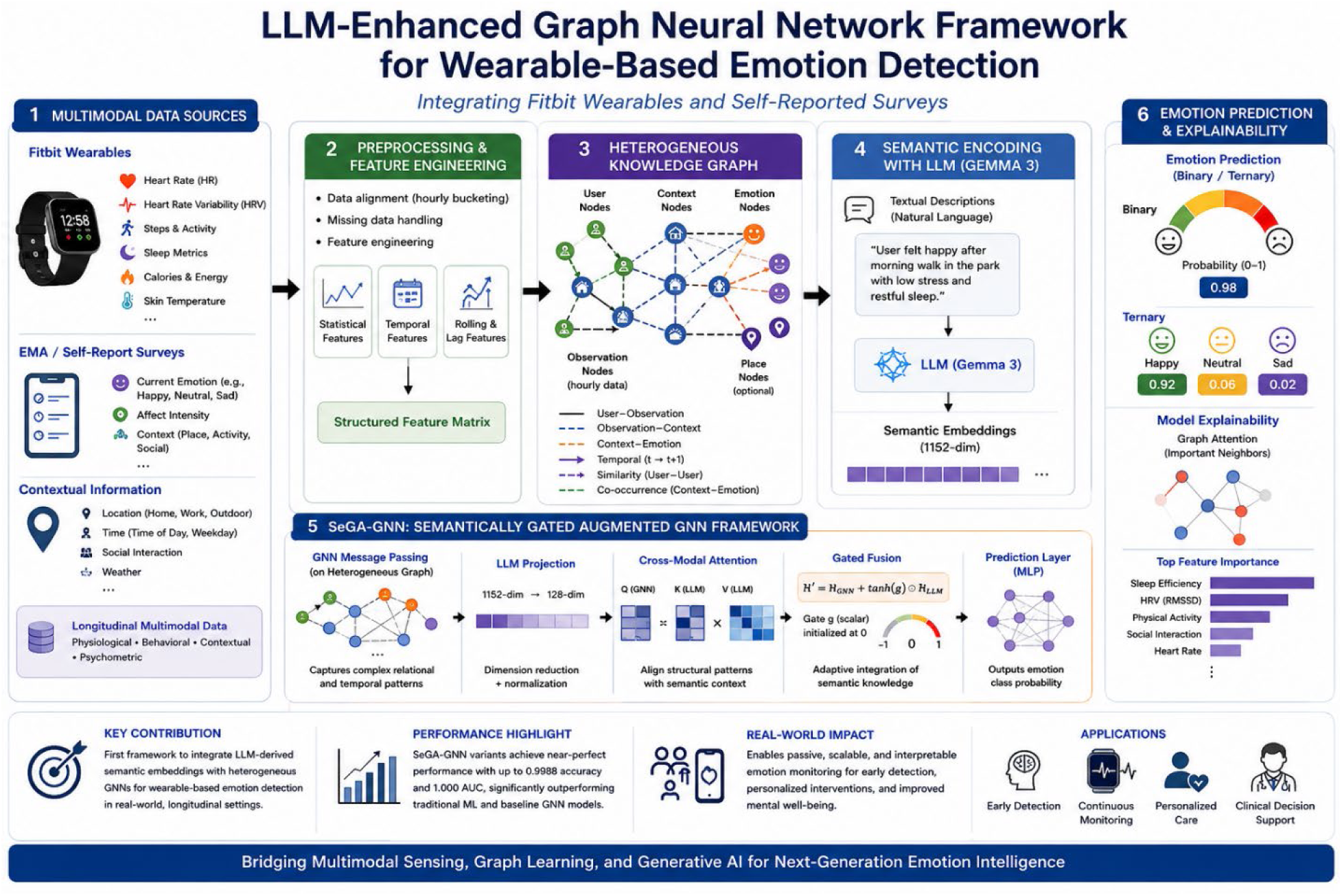

## 1. Introduction

For millions of people worldwide, migration represents not only a geographic transition but a significant psychological toll, with first-generation migrants showing consistently elevated rates of common mental disorders relative to settled populations [1]. Among refugees, rates of post-traumatic stress disorder are substantially higher than those observed in the general population, reflecting the cumulative weight of pre-departure trauma, forced displacement, and post-arrival adversity [1]. This vulnerability is not the product of a single traumatic event; it accumulates across the entire trajectory of displacement, from exposure to conflict and violence before departure, through the hardships of the journey, to the legal uncertainty and social isolation that often persist well into resettlement [2]. What makes this particularly consequential from a public health perspective is that the very mechanisms underlying this burden (loss of control, unmet basic needs, and social disconnection) are precisely those that impede meaningful integration into host societies [3].

Addressing mental health needs at this scale demands monitoring approaches that extend beyond the clinic. Passively collected data from smartphones and wearable devices, spanning physical activity, sleep, mobility, and digital behavior, have been shown to carry meaningful signals of psychological state, with classification accuracies exceeding 80% in controlled settings [4], [5]. These signals emerge naturally from how individuals move through and interact with their daily environment, capturing behavioral variability without placing additional demands on already burdened populations. Yet, the current landscape of digital phenotyping and passive sensing exhibits a pronounced demographic and geographic bias [6]. Existing literature has largely focused on homogeneous, low-risk cohorts, such as college students [7], [8], alongside individuals embedded in urban, high-resource, or stable clinical environments [9]. Because current predictive algorithms are frequently trained on these specific Western, high-income cohorts, they inherently carry algorithmic bias and lack generalizability to culturally distinct or low-resource settings [9], [10]. Consequently, communities carrying the heaviest psychological burden, among them migrants and refugees, remain almost entirely absent from this research. This systemic exclusion leaves a critical gap between where these scalable monitoring tools are currently developed and where they are most urgently needed.

## 2. Background/Literature Review

Affective computing, broadly understood as the design of systems capable of recognizing, interpreting, and responding to human emotional states, emerged as a formal discipline with Picard’s foundational framing [11] and has since expanded into a broad research agenda spanning facial analysis, physiological monitoring, natural language processing, and multimodal sensor fusion [12]. A defining development within this trajectory has been the convergence of machine learning with ubiquitous sensing: passively collected data from smartphones and wearable devices now offer continuous, low-burden windows into behavioral and psychological states that were previously accessible only through scheduled clinical contact [13]. Doryab and colleagues demonstrated that combined smartphone and Fitbit data could classify loneliness levels with 80.2% accuracy from activity, mobility, and sleep features alone [4], while Liang and colleagues showed that multimodal mobile behavior remains predictive of daily mood even in high-risk populations [5]. Despite these advances, populations navigating sustained adversity, including migrants and refugees, remain profoundly underrepresented in the digital mental health literature [14]. Even when these highly vulnerable groups are included in existing research, studies frequently suffer from small sample sizes and significant selection biases, such as the overrepresentation of highly educated individuals or specific demographic subsets, which severely limits the generalizability of the findings [15], [16]. Consequently, translating scalable passive sensing technologies from controlled environments to the complex, real-world conditions of forced displacement and psychological vulnerability remain a critical and unresolved gap in the literature [14], [17].

Affective computing has traditionally explored single-modality approaches, with text-based and visual methods representing two of the most extensively studied directions. In textual analysis, Mahima and colleagues argued that sentiment analysis alone is insufficient for capturing the complexity of human emotional expression, proposing a hybrid architecture combining BERT-based sentence embeddings with contextual rules to enable multi-label detection across Ekman’s basic emotion categories [18]. Cahyani and colleagues confirmed the advantage of transformer-based embeddings empirically, showing that a BERT and CNN combination achieves 86–87% accuracy on text corpora, outperforming Word2Vec and GloVe baselines [19]. Deng and Ren extended this further with an architecture that explicitly models the dependencies between simultaneously expressed emotions, a capability that single-label classifiers structurally cannot provide [20]. Parallel to these textual advancements, facial expression recognition has attracted the largest volume of work within the visual domain. Here, convolutional and attention-based architectures have achieved strong performance under controlled conditions across contexts ranging from classroom monitoring to elderly care [21], [22], [23], [24], [25], [26], [27].

Expanding beyond text and vision, research in the auditory domain has increasingly shifted toward temporally resolved emotion tracking. Yousef and colleagues proposed a dual-stage system combining LSTM networks with the PELT change point detection algorithm, achieving 91.04% accuracy on discrete emotion identification and 89.43% transition detection accuracy at a two-second tolerance [28]. Wang and colleagues formalized this as Speech Emotion Diarization, framing the task as determining which emotion appears when rather than assigning a single label to an entire clip [29]. Complementing these external behavioral cues, physiological signals, particularly EEG, offer a more objective window that operates independently of language and cultural background. These signals connect naturally to the arousal-valence framework that has informed our own valence-based classification scheme [30].

However, the limitations of relying on isolated signals have driven the field toward multimodal integration. Lee and colleagues provide direct empirical support for this shift. Their ablation analyses demonstrate that audio and visual components each make independent contributions to recognition performance when combined with BERT-derived linguistic representations, with attention-based fusion mechanisms consistently outperforming simple feature concatenation [6]. Jabeen and colleagues confirm this pattern across a broader modality spectrum spanning image, video, text, audio, and physiological signals. Yet, they also document a persistent challenge: learning meaningful cross-modal associations, rather than merely pooling modality-specific predictions, remains an open problem [31]. A consequential development in addressing these cross-modal challenges is the explicit incorporation of temporal structure into multimodal frameworks. Li and colleagues argue that human emotions are better understood as continuously evolving streams than as static labels attached to well-trimmed clips. Their AVES dataset annotates videos with precise temporal boundaries of emotion segments rather than a single category per clip, and their Boundary Combination Network exploits short-term temporal context to first locate these boundaries before classifying the contained emotions [32].

Recent literature confirms that traditional multimodal fusion strategies, which typically project signals into a shared latent space or concatenate features, yield suboptimal results by failing to concurrently capture modality-specific heterogeneity and intrinsic cross-modal relationships [33], [34]. Furthermore, conventional architectures struggle with the inherent ambiguity of knowledge transfer between behavioral and physiological signals [35] and cannot effectively encode the higher-order, many-to-many interactions present in demographic and social data [36]. Consequently, mapping these multi-modal dependencies explicitly via structural graph topologies has become a necessity. To address this structural challenge, Kipf and Welling established that convolutional filters operating on graph-structured data can learn node representations encoding both local structural context and node-level features [37]. Hamilton and colleagues extended this to an inductive setting through GraphSAGE, learning a neighborhood aggregation function that generalizes to nodes unseen during training [38]. Both architectures, however, assume homogeneous graphs in which all nodes and edges belong to the same type, an assumption that rarely holds in multimodal behavioral data. The Heterogeneous Graph Transformer addressed this by introducing type-dependent parameter matrices that maintain distinct representations for structurally different entity types [39], while Wang and colleagues complemented this with a hierarchical attention mechanism that learns the importance of both individual neighbors and entire meta-paths, enabling richer semantic reasoning over heterogeneous topologies [40].

Formalizing these heterogeneous relationships as a knowledge graph further extends the model’s capacity to exploit structural dependencies. Nickel and colleagues demonstrated that statistical models trained on knowledge graphs can generalize beyond observed relationships through latent feature decomposition and observable pattern mining [41]. Furthermore, Rotmensch and colleagues showed that automated knowledge graph construction from medical records achieves a precision of 0.85 against expert physician evaluations, establishing the feasibility of this approach in health informatics [42]. Crucially, enriching these structural representations with semantic context from Large Language Models (LLMs) bridges the gap between structured relational data and unstructured behavioral cues. Pre-trained transformer architectures demonstrate that deep representations learned from unlabeled text transfer effectively to a wide range of downstream tasks [43], making them attractive as feature encoders where structured data falls short. Hwang and colleagues demonstrated the viability of this combination, showing that fusing LLM-derived embeddings with GNN representations captures subtle behavioral patterns that neither modality detects alone [44].

Building upon these foundations, our study applies this principle to affective state prediction. By encoding natural language descriptions of physiological observations and user profiles through a frozen instruction-tuned language model, we inject the resulting semantic embeddings into the GNN, successfully incorporating contextual meaning that raw sensor features cannot inherently express.

## 3. Materials and Methods

In this section, we detail the comprehensive methodology developed to predict user affective states by integrating longitudinal physiological signals with self-reported psychological surveys. The proposed analytical pipeline is structured into several progressive phases, transitioning from raw data ingestion to the deployment of advanced neural architectures. First, we describe the raw data sources and the rigorous data cleaning and feature engineering protocols applied to ensure high-quality, reliable inputs. Following data preparation, we outline the construction of a heterogeneous knowledge graph, specifically designed to capture the complex, multi-relational, and temporal dynamics among users, behavioral observations, contextual settings, and mood states. To rigorously evaluate our approach, we establish a two-tiered baseline: traditional tabular machine learning models and a suite of state-of-the-art heterogeneous Graph Neural Networks (GNNs). Finally, we introduce the core architectural contribution of this study: an LLM-enhanced GNN framework that leverages a foundational language model (Gemma 3 [45]) to inject deep semantic contextualization into the graph representations via a gated cross-modal fusion mechanism. The following subsections sequentially describe each phase of this experimental design.

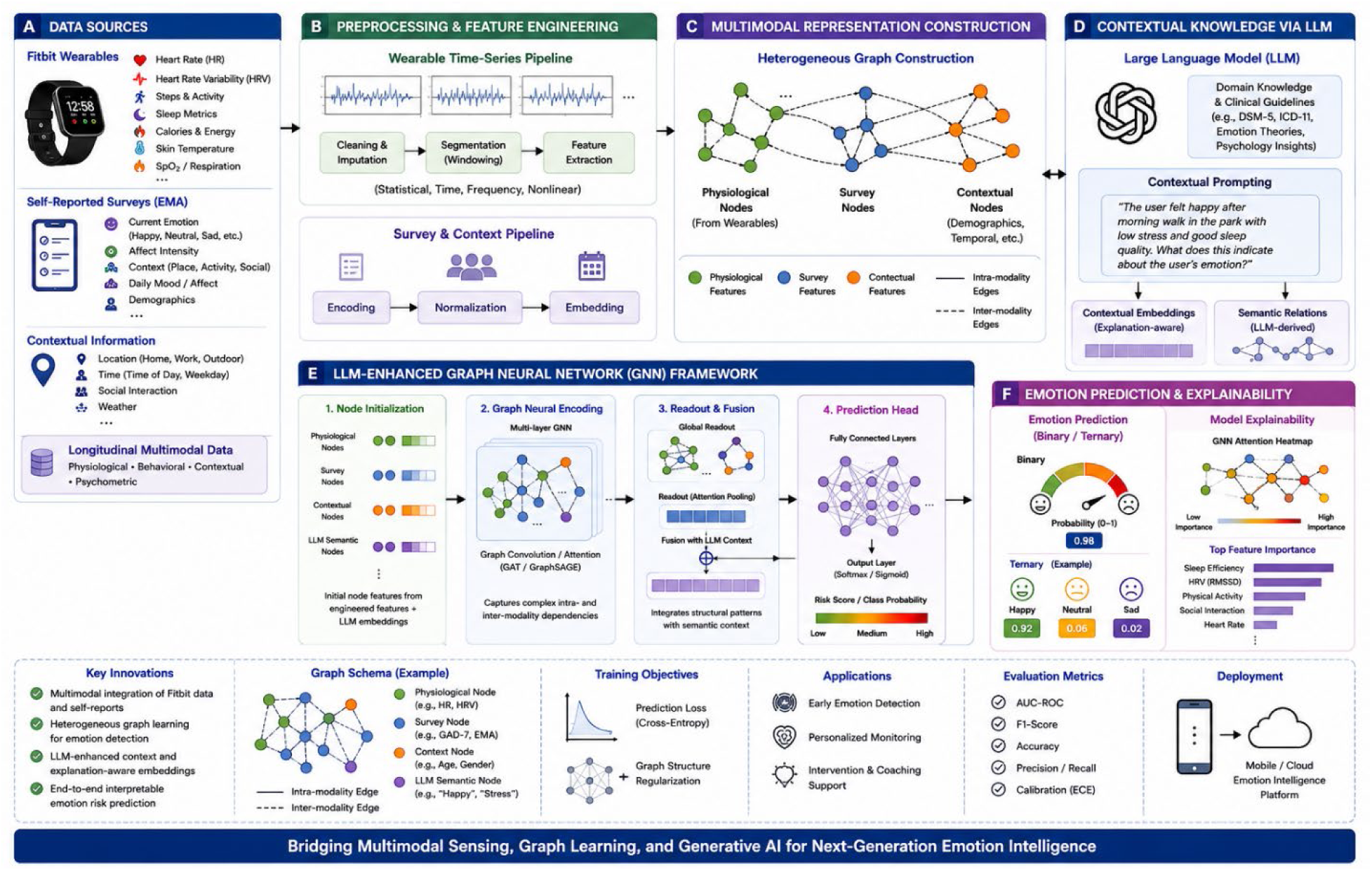

### 3.1. Data

The empirical foundation of this study is the LifeSnaps [46] dataset, a large-scale, multi-modal, and longitudinal open-source dataset. It captures the daily lives of participants over an uninterrupted period of four months, providing a high-resolution view of the interplay between objective physiological signals and subjective psychological states. The dataset originally included data from 71 participants, recruited to reflect a diverse range of demographic backgrounds. The data collection architecture is centered around a multi-modal approach:

- **Physiological and Biometric Stream:** High-frequency data was captured via Fitbit Charge 2 and Charge 3 wearable devices. This stream provides second-by-second heart rate data, minute-level step counts, and energy expenditure (calories). Furthermore, the dataset contains daily sleep logs that decompose sleep into specific stages (Light, Deep, REM, and Awake) alongside metrics for sleep efficiency and duration. It also includes daily summaries of heart rate variability (HRV) and nightly skin temperature variations, offering a holistic view of the users’ autonomic nervous system activity.
- **Psychological and Contextual Stream:** Subjective data was collected using the SEMA (Smartphone Ecological Momentary Assessment) application. Participants responded to several prompts per day, yielding a rich longitudinal record of their affective states across multiple dimensions (e.g., valence, arousal, and specific emotions like happiness, tension, or fatigue). In addition to mood, the SEMA surveys captured the social and environmental context (e.g., location, current activity, and social company), providing the necessary ground truth for contextualizing the physiological signals.
- **Structured Surveys:** Beyond momentary assessments, the dataset includes validated psychometric instruments administered at baseline and follow-up, such as the Positive and Negative Affect Schedule (PANAS) and State-Trait Anxiety Inventory (STAI). These provide stable psychological profiles for each user, which we utilize to enrich the node features within our graph-based models.

The raw data is organized in a MongoDB database comprising over 71 million biometric records and thousands of survey responses. This scale allows for the exploration of complex temporal patterns and the development of robust machine learning models capable of generalizing different user behaviors.

### 3.2. Data Preprocessing and Cleaning

The raw data extracted from MongoDB underwent a rigorous multi-stage preprocessing pipeline to transform high-frequency, multi-modal signals into a structured, model-ready format. This pipeline was designed to handle the inherent sparsity of wearable data while preserving the temporal integrity of effective self-reports.

#### Data Alignment and Hourly Bucketing

Given the asynchronous nature of Fitbit biometrics and SEMA surveys, we established an hourly temporal grid as the primary unit of observation. This granularity was justified by an initial exploration of the SEMA response frequency, which revealed a median gap of approximately 90 minutes between entries, making hourly bucketing a stable window for capturing contextual shifts without excessive data loss. For instances where multiple labels occurred within the same hour, a “last-observed” strategy was applied to prioritize the most proximal affective state.

#### Label Cleaning and Transformation

The target variable was derived from SEMA surveys by filtering for completed responses and removing entries with missing mood data. To facilitate binary classification, the original multi-class affective states were mapped into a valence-based binary scale: Positive (*comprising Rested/Relaxed, Happy, and Alert*) and Negative (*comprising Tense/Anxious, Sad, and Tired*). Observations labeled as *Neutral* or those with ambiguous responses were excluded to ensure clear class separability.

#### Feature Engineering and Temporal Encoding

To capture the dynamic nature of physiological states, we performed extensive feature engineering and temporal encoding. High-frequency Fitbit signals; specifically, heart rate, steps, calories, and distance were first aggregated into comprehensive hourly statistics, yielding mean, minimum, maximum, and standard deviation metrics for each temporal window. To preserve the cyclical continuity of time, periodic variables such as the hour of the day and the day of the week were mathematically transformed using sine and cosine functions.

Furthermore, to account physiological memory and behavioral inertia, we computed rolling averages over 3-, 6-, and 24-hour windows for key biometric signals, alongside incorporating lagged affective labels to capture individual mood persistence. Finally, missing data was addressed through a dual management strategy: a limited forward-fill mechanism, restricted to a maximum of three hours, was applied to maintain the temporal flow of high-frequency signals. This was concurrently supplemented by the generation of binary indicator flags to explicitly denote these imputed intervals to the downstream predictive models.

#### Statistical Quality Control

The final cleaning stage involved fine-grained filtering to ensure data reliability. We removed participants with insufficient longitudinal coverage, settling on a final cohort of 62 users. Outlier detection was performed using the Interquartile Range (IQR) method for physical activity metrics. Furthermore, heart rate observations (BPM) were nullified for hours where the sample count was too low to be statistically representative. Final missing values were imputed using a user-specific median strategy, ensuring that the personal baseline of each participant was respected.

The culmination of this preprocessing pipeline is a refined dataset comprising over 4,100 high-quality hourly observations from 62 unique participants. This dataset serves a dual purpose in our methodology: first, it provides the structured feature matrix and target labels required to train and evaluate our baseline machine learning models. Second, it acts as the primary substrate for constructing the heterogeneous knowledge graph, where each hourly record is instantiated as an *Observation* node, enriched with the engineered physiological and temporal features. By establishing this consistent data foundation, we ensure a fair and rigorous comparison between traditional tabular approaches and graph-based architecture.

### 3.3. Knowledge Graph Construction

To fully capture the complex temporal and relational dependencies inherent in longitudinal behavioral data, we modeled the preprocessed dataset as a heterogeneous knowledge graph 𝒢𝒢 = (𝒱𝒱, ℰ). The construction of this graph was carried out in two phases: an initial exploratory data analysis to assess cohort viability and feature alignment, followed by the systematic generation of a high-density, learnable graph topology.

#### 3.3.1. Cohort Selection and Node Initialization

Based on our exploratory analysis of the cleaned hourly data, we restricted the graph to a robust sub-cohort of 45 users (comprising 4,030 observation windows) who exhibited high overlap (≥80%) with the baseline psychometric surveys. This ensured sufficient data density for user profiling. The resulting heterogeneous graph comprises four distinct node types (𝒱𝒱):

- **Observation Nodes** (𝑽_𝒐bs_ 𝒏 = 𝟒,**030**): Representing hourly behavioral snapshots, each node is initialized with a 61-dimensional feature vector containing standardized physiological metrics and temporal encodings.
- **User Nodes** (𝑽*_user_*, 𝒏 = 𝟒5): Representing the participants. Missing survey responses were imputed to create a complete 61-dimensional psychometric and aggregated baseline profile for each user, allowing the graph to model stable individual traits.
- **Mood Context Nodes** (𝑽_𝒎ood_, 𝒏 = 𝟔): Representing the distinct affective states (e.g., ALERT, HAPPY, TENSE/ANXIOUS). These are encoded with a 10-dimensional feature vector combining semantic valence/arousal representations and one-hot encodings.
- **Place Context Nodes** (𝑽*_place_*, 𝒏 = 𝟖): Representing physical locations (e.g., HOME, WORK, TRANSIT), encoded via a 14-dimensional vector of semantic and one-hot features.

#### 3.3.2. Edge Topology and Relational Dynamics

To maximize the communicative capacity of the graph, we engineered a sophisticated connectivity schema ℰ resulting in approximately 67,600 edges across distinct relational types:

- **User-Observation Edges (Ownership):** Bipartite edges (*has_observation* and *belongs_to*) connect each of the 4,030 observations to their respective user nodes.
- **User-User Edges (Homophily):** Initial EDA revealed that simple threshold-based similarity resulted in isolated users. Therefore, we connected users based on a hybrid approach: a k-Nearest Neighbors (k=5) graph computed over their psychophysiological profiles, augmented by temporal co-activity edges for users sharing ≥ 3 hours of concurrent data. This yielded 1,060 *similar_to* connections.
- **Temporal Edges (Sequential Flow):** To capture affective inertia, directed edges were constructed between consecutive observations. This included both within-day sequences and cross-day bridges (connecting the last observation of one day to the first of the next), resulting in 3,985 forward (*next_obs*) and 3,985 reverse (*prev_obs*) edges.
- **Observation-Observation Edges (State Similarity):** To enable the model to generalize across different participants experiencing similar physiological states, we computed a cross-user feature-space k- Nearest Neighbors (k=10), generating 38,506 *similar_obs* edges.
- **Contextual Edges:** Observations were linked to their concurrent environmental context via 4,029 *at_place* and corresponding *rev_at_place* edges, as well as 4,030 *has_mood* edges.

#### 3.3.3. Strict Leakage Prevention

Because the target variable is a binary classification of the mood state, the *mood_label* is highly deterministic. To prevent the GNN models from exploiting structural shortcuts during message passing, rigorous leakage control mechanisms were enforced. Specifically, reverse mood-to-observation edges (*rev_has_mood*), mood-to-mood transition edges, and mood-place co-occurrence edges were completely excluded from the graph. Furthermore, when computing the cross-user observation similarity edges (*similar_obs*), 6 label-adjacent features (such as previous affective labels and longitudinally aligned PANAS/STAI scores) were masked out, restricting the k-NN algorithm to only 55 purely physiological and temporal dimensions.

### 3.4. Baseline Heterogeneous GNN Models

To effectively capture the multi-relational and temporal dynamics structured within our knowledge graph (𝒢𝒢), we formulated the predictive task as a binary node classification problem on the observation nodes (V_obs_). Unlike the tabular baselines that treat each observation in isolation, GNNs leverage message passing to aggregate physiological, psychological, and contextual information from a node’s local neighborhood. To establish a rigorous foundation before introducing our novel Large Language Model (LLM) enhancements, we evaluated five state-of-the-art heterogeneous GNN architectures.

#### 3.4.1. HeteroSAGE

Operationally, the HeteroSAGE [38] model is structured as a two-layer stacked neural network. In each layer, a HeteroConv wrapper dictates the message passing by applying SAGEConv [38] operators across all edge types. For each relation, neighbors are aggregated using a mean-pooling strategy, and the resulting relation-specific messages are fused at the target node via sum-aggregation. To stabilize the internal covariate shift across the highly varied multi-modal features, a 1D Batch Normalization (BatchNorm) layer is applied exclusively to the observation node representations immediately following the convolutional step. This normalized output is then processed through a ReLU activation function and a dropout layer (p=0.3) before being fed into the second message-passing layer. Finally, the refined 128-dimensional embeddings of the observation nodes are pass

#### 3.4.2. Heterogeneous Graph Attention Network (HeteroGAT)

The Heterogeneous Graph Attention Network [47] maps input features through two sequential attention-based convolutional layers. Within the HeteroConv framework, a GATConv [47] module calculates normalized self-attention scores for neighbors across every specific relation. To capture diverse structural patterns without causing dimensional explosion, the architecture utilizes 4 independent attention heads per layer. Crucially, the outputs of these multiple heads are averaged rather than concatenated, ensuring the hidden representation strictly maintains its 128-dimensional size. Self-loops are explicitly disabled within the attention operator, relying on the heterogeneous wrapper to manage the central node’s residual updates. Following two iterations of attention, ReLU activation, and dropout, the final observation embeddings are linearly projected to the target space.

#### 3.4.3. Heterogeneous Graph Transformer (HGT)

The HGT [39] architecture natively circumvents the need for a generic heterogeneous wrapper by utilizing two layers of HGTConv [39]. Instead of shared weight matrices, this architecture assigns distinct, type-specific parameter matrices for different node and edge types. Operating within a 128-dimensional hidden space configured with 4 attention heads, each head computes mutual attention in a 32-dimensional subspace. During the forward pass, type-specific linear projections generate the necessary Query, Key, and Value matrices. The model computes an attention-weighted message aggregation, applies a residual connection, and follows with a ReLU activation and 0.3 dropout. After two complete Transformer-like graph propagation steps, a fully connected linear layer acts as the readout mechanism for the observation nodes.

#### 3.4.4. Heterogeneous Graph Attention Network (HAN)

The HAN [40] model is constructed using two stacked HANConv [40] stages to perform hierarchical attention. In the first layer, raw node features are mapped into a shared 128-dimensional latent space. The model then executes node-level attention, aggregating features from neighboring nodes along explicitly defined semantic metapaths. This is immediately followed by a semantic-level attention mechanism, which learns optimal weights to fuse these distinct metapath-specific embeddings into a unified representation. This dual-level attention output is passed through a ReLU activation and dropout layer before entering the second HANConv stage for further topological refinement. The terminal 128-dimensional representations of the observation nodes are then passed to the linear classifier.

#### 3.4.5. Simple and Efficient Heterogeneous Graph Neural Network (SeHGNN)

Unlike standard layer-by-layer message passing on a heterogeneous graph, the SeHGNN-style [48] module follows the idea of Yang et al. by working in a single homogeneous index space over all node types, with row-normalized adjacency operators per relation. Node features are first mapped to a common hidden size with a shallow nonlinear projection. For each metapath template, features are updated by mean aggregation along that template using products of these fixed operators; one channel uses no extra propagation beyond the base representation. The templates encode forward and backward temporal order, similarity-based ties between target entities, two-hop context centered on an actor type, and context mediated by an auxiliary spatial or environmental type with a return path. Only rows corresponding to the target entity type for the downstream task are retained from each channel. The resulting channels form a sequence processed by a two-layer Transformer encoder with four attention heads per layer, feedforward width four times the hidden dimension, and dropout 0.3, so self-attention fuses relational channels instead of aggregating only one-hop neighbors. Channel outputs are mean-pooled along the sequence into a 128-dimensional embedding per target entity, followed by a linear classifier to produce logits.

### 3.5. The SeGA-GNN Framework

In this section, we introduce the **Semantically Gated Augmented GNN (SeGA-GNN)** framework, designed specifically to isolate and observe the direct impact of Large Language Models on predictive performance. To rigorously evaluate this effect and understand how semantic augmentation influences the results, we intentionally preserve the exact architectural configurations of the baseline heterogeneous GNNs. By keeping the underlying message-passing structures identical and augmenting them strictly through our proposed cross-modal fusion mechanism, SeGA-GNN ensures that any improvements in classification outcomes are directly attributable to the LLM integration. Rather than performing computationally prohibitive joint finetuning, SeGA-GNN utilizes a two-stage pipeline that injects rich, pre-computed semantic knowledge from an instruction-tuned foundational model into the established graph topology. The fusion block was systematically integrated into all five baseline architectures, resulting in **SeGA-HeteroSAGE**, **SeGA-HeteroGAT**, **SeGA-HGT**, **SeGA-HAN**, and **SeGA-SeHGNN**.

#### 3.5.1. Offline Semantic Encoding

In the first phase of SeGA-GNN, we generated descriptive natural-language strings for every node in the graph (𝒱𝒱). For observation nodes, this text synthesized the hourly biometric aggregates; for user nodes, it articulated their psychological profiles; and for context nodes, it defined the semantic meaning of specific moods and locations. To encode these textual representations, we utilized **Google’s Gemma 3 (1B IT)** [45], an advanced instruction-tuned foundational model. Operating in a frozen, offline state (fp16 precision), the LLM processed the text batches. To extract a unified semantic vector for each node, we isolated the *last_hidden_state* of the model, applied attention-masked mean-pooling to aggregate the sequence into a single representation, and finally applied row-wise L2-normalization. This procedure yielded dense, 1152-dimensional semantic embeddings for each node, which were saved and stored independently, allowing the massive LLM to be completely offloaded from GPU memory prior to GNN training.

#### 3.5.2. Gated Cross-Modal Fusion Mechanism

The *Gated* component of SeGA-GNN is implemented through a custom LLM Fusion Block, which is injected immediately following every message-passing layer in the backbone architectures. The fusion process operates through three sequential steps for every node type:

- **Dimensionality Reduction:** A linear projection layer compresses the frozen 1152-dimensional LLM embedding down to the 128-dimensional hidden space of the GNN.
- **Cross-Modal Attention:** A Multi-Head Attention mechanism (configured with 4 heads) is applied. In this single-step cross-attention operation, the dynamically evolving GNN hidden state acts as the *Query*, while the projected LLM embedding serves as both the *Key* and *Value*. This allows the structural node representation to actively attend to its relevant semantic context.
- **Zero-Initialized Gating:** The attended semantic vector is fused back into the GNN state using a learnable residual gate and Layer Normalization:

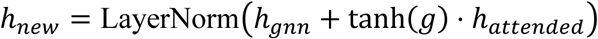

Crucially, the learnable scalar parameter 𝒈𝒈 (the gate) is initialized to strictly zero. At epoch zero, tanh(0) = 0, meaning the fusion branch is entirely inactive and the model behaves exactly like a pure GNN. As training progresses, SeGA-GNN dynamically learns to open this gate, smoothly injecting semantic information only when it minimizes classification loss.

## 4. Results and Discussion

### 4.1 . Experimental Setup

To comprehensively evaluate the effectiveness of the proposed framework, we assessed two categories of graph-based predictive models. The first category consisted of baseline heterogeneous Graph Neural Network (GNN) architectures, including HeteroSAGE, HeteroGAT, HGT, HAN, and SeHGNN, which were trained directly on the constructed heterogeneous knowledge graph. The second category comprised the proposed Semantically Gated Augmented Graph Neural Network (SeGA-GNN) variants, where the baseline GNN architectures were enhanced with semantic embeddings generated from a Large Language Model (LLM) through the proposed gated cross-modal fusion mechanism. This evaluation strategy enabled a systematic comparison between conventional heterogeneous graph learning and semantically augmented graph learning approaches.

#### 4.1.1. Data Partitioning

To ensure rigorous evaluation and prevent information leakage, a strict per-user temporal splitting strategy was employed for all graph-based models. Specifically, the observation nodes were partitioned chronologically into 70% training, 10% validation, and 20% testing subsets on a per-user basis. This temporal evaluation setup preserves chronological integrity and prevents future-data leakage by ensuring that future observations are never used during model training. Such a strategy more accurately reflects realistic longitudinal forecasting scenarios in digital mental health applications, where future affective states must be predicted using only historical physiological and contextual information.

#### 4.1.2. Training Protocol and Hyperparameters

All graph-based models, including both baseline heterogeneous GNN architectures and the proposed SeGA-GNN variants, were trained using a unified full-batch training configuration to ensure fair comparison across architectures. Optimization was performed using the Adam optimizer with a learning rate of 1 × 10^−3^and a weight decay coefficient of 1 × 10^−4^. To address potential class imbalance in the anxiety classification task, a class-weighted Cross-Entropy Loss function was employed during training.

The graph architectures were configured using two message-passing layers with a 128-dimensional hidden representation space and a dropout rate of 0.3 to reduce overfitting. Attention-based architectures, including HGT, HAN, and the SeGA-enhanced models, utilized four attention heads to improve the modeling of heterogeneous relationships and contextual dependencies.

For the proposed SeGA-GNN framework, semantic node embeddings were generated offline using a frozen Gemma 3 (1B IT) Large Language Model. During training, gradient clipping with a threshold of 1.0 was applied to improve numerical stability and prevent gradient explosion during optimization. All models were trained for a maximum of 200 epochs, and the final model checkpoint was selected based on the highest validation ROC-AUC score to ensure optimal generalization performance.

#### 4.1.3. Implementation

All experiments were implemented using the PyTorch Geometric framework and executed on NVIDIA A100 GPUs. The graph-based models were developed using PyTorch Geometric to efficiently support heterogeneous graph construction and message-passing operations. This unified implementation environment enabled reproducible experimentation and efficient large-scale training of both baseline heterogeneous GNN architectures and the proposed semantically enhanced SeGA-GNN models.

### 4.2. Performance Evaluation measures

To comprehensively evaluate the effectiveness of the proposed **SeGA-GNN framework** and the baseline heterogeneous Graph Neural Network architectures, multiple classification performance metrics were employed. Since wearable-based emotion detection involves both **binary and ternary classification scenarios** for recognizing different emotional states, the evaluation protocol was designed to assess not only overall predictive performance but also the balance between class discrimination capability, sensitivity to emotional variations, and classification robustness across multiple affective categories.

The models were evaluated using **Accuracy, Precision, Recall, F1-score, and Area Under the Receiver Operating Characteristic Curve (AUC-ROC)**. Accuracy was used to quantify the overall proportion of correctly classified emotional states. Precision measured the reliability of positive emotional predictions, while Recall evaluated the ability of the models to correctly identify target emotional categories. The F1-score was employed to provide a balanced assessment of Precision and Recall, particularly under potentially imbalanced emotional class distributions. Additionally, AUC-ROC was used to evaluate the discriminative capability of the models across different decision thresholds and to assess the robustness of classification performance.

Together, these evaluation measures provide a comprehensive assessment of model effectiveness by simultaneously capturing overall predictive accuracy, class separability, sensitivity to emotional states, and the ability of the proposed framework to generalize across heterogeneous multimodal wearable sensing environments.

Accuracy measures the proportion of correctly classified observations among all samples and is defined as:

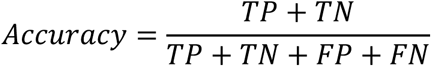

where 𝑇*P*, 𝑇*N*, 𝐹*P*, and 𝐹*N* denote true positives, true negatives, false positives, and false negatives, respectively. Although accuracy provides an overall measure of classification performance, it may not fully reflect model behavior in cases of class imbalance.

Precision evaluates the proportion of correctly predicted positive samples among all samples predicted as positive and is calculated as:

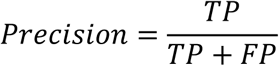

High precision indicates that the model generates fewer false positive predictions, which is particularly important in mental health applications to avoid unnecessary anxiety alerts or interventions.

Recall, also referred to as sensitivity, measures the proportion of actual positive samples that are correctly identified by the model:

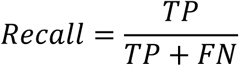

Recall is especially important in anxiety detection tasks because failing to identify true anxiety-related states may lead to missed opportunities for early intervention and monitoring.

The F1-score provides a harmonic balance between precision and recall and is defined as:

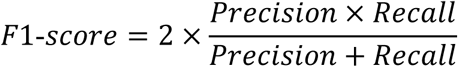

This metric is particularly useful when evaluating models under potentially imbalanced class distributions, as it simultaneously accounts for both false positives and false negatives.

To further assess the discriminative capability of the models across different classification thresholds, the Area Under the Receiver Operating Characteristic Curve (AUC-ROC) was computed. The ROC curve represents the trade-off between the True Positive Rate (TPR) and False Positive Rate (FPR), where:

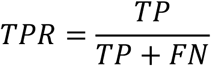

and

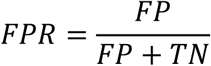

The AUC value summarizes the overall separability between positive and negative classes. A higher AUC indicates stronger classification capability and greater robustness across varying decision thresholds.

Together, these evaluation metrics provide a comprehensive assessment of model performance, enabling detailed analysis of predictive accuracy, class discrimination capability, sensitivity to anxiety-related states, and robustness of the proposed SeGA-GNN framework compared to baseline heterogeneous GNN architectures.

### 4.3. Performance Analysis of Baseline Heterogeneous GNN Models

Table 1 presents the binary classification performance of five heterogeneous Graph Neural Network (GNN) architectures—HeteroSAGE, HeteroGAT, HGT, HAN, and SeHGNN—trained on the constructed heterogeneous knowledge graph for **emotion detection**. The evaluation was performed using Accuracy, Precision, Recall, F1-score, and AUC to comprehensively assess predictive capability, classification balance, and discriminative performance across emotional states.

**Table 1:** The Binary Classification Performance of Baseline GNN Models - Trained with the Knowledge Graph.

| **Baseline GNN Model** | **Accuracy** | **Precision** | **Recall** | **F1-Score** | **AUC** |
| --- | --- | --- | --- | --- | --- |
| **HeteroSAGE** | 0.9677 | **0.9685** | 0.9663 | 0.9676 | **0.9985** |
| **HeteroGAT** | 0.9502 | 0.9523 | 0.9825 | 0.9502 | 0.9946 |
| **HGT** | **0.9739** | 0.9748 | 0.9814 | **0.9739** | 0.9978 |
| **HAN** | 0.7525 | 0.7536 | **0.9988** | 0.7523 | 0.8483 |
| SeHGNN | 0.9167 | 0.9169 | 0.9116 | 0.9166 | 0.9576 |

Overall, the results demonstrate that heterogeneous graph-based learning provides highly effective predictive performance for multimodal emotion detection. Most models achieved exceptionally high classification accuracy and AUC values approaching 1.0, indicating that the knowledge graph representation successfully captures complex interactions among physiological signals, behavioral patterns, contextual information, and user-specific temporal relationships that contribute to emotional state recognition.

Among the evaluated architectures, HGT achieved the strongest overall performance, obtaining the highest accuracy (0.9739), precision (0.9748), F1-score (0.9739), and a near-perfect AUC of 0.9978. Additionally, HGT achieved the highest recall among the top-performing models (0.9814), indicating superior sensitivity for identifying emotional states. These findings suggest that transformer-based heterogeneous graph architectures are particularly effective for multimodal emotion modeling. The type-aware attention mechanism employed by HGT enables selective information aggregation across different node and edge categories, allowing the model to capture complex relational dependencies while preserving multimodal heterogeneity.

HeteroSAGE demonstrated similarly strong and highly balanced performance, achieving an accuracy of 0.9677, precision of 0.9685, recall of 0.9663, and an F1-score of 0.9676, together with the highest AUC value among all baseline models (0.9985). The close agreement among all evaluation metrics suggests highly stable classification behavior with minimal bias toward any emotional category. This indicates that neighborhood aggregation through mean-based message propagation is highly effective in preserving local structural information within the heterogeneous graph. The results suggest that even relatively simple aggregation strategies can effectively exploit multimodal emotional relationships when supported by an appropriately designed graph structure.

HeteroGAT also achieved strong performance, reaching an accuracy of 0.9502 and an AUC of 0.9946. A notable characteristic of HeteroGAT is its exceptionally high recall (0.9825), which exceeds its precision (0.9523). This behavior indicates that the model is particularly effective at identifying target emotional states, although it may generate slightly more false positive predictions compared to HGT and HeteroSAGE. Such behavior is consistent with attention-based architectures that dynamically assign importance to neighboring nodes and may preferentially emphasize strongly informative emotional patterns. In wearable-based emotion detection, high recall is valuable because failing to recognize relevant emotional conditions may reduce responsiveness and personalization of emotion-aware systems.

In contrast, HAN exhibited substantially lower overall performance, achieving an accuracy of 0.7525 and an AUC of 0.8483 despite producing an extremely high recall of 0.9988. Although this recall suggests excellent sensitivity to emotional transitions, the substantially lower precision (0.7536) and F1-score (0.7523) indicate excessive false positive predictions. This imbalance suggests that the hierarchical attention mechanism employed by HAN may overemphasize dominant metapath relationships and amplify specific semantic connections, reducing the model’s ability to generalize across diverse emotional patterns. The findings indicate that hierarchical attention alone may be insufficient for effectively modeling heterogeneous multimodal emotion data without additional contextual regularization.

SeHGNN achieved moderate but stable performance, obtaining an accuracy of 0.9167 and an AUC of 0.9576. Precision (0.9169), recall (0.9116), and F1-score (0.9166) remained highly balanced, reflecting consistent classification behavior. The performance of SeHGNN suggests that semantic aggregation through precomputed metapath propagation provides efficient representation learning but may not fully exploit dynamic interactions among heterogeneous entities. Consequently, while SeHGNN demonstrates strong performance, it does not reach the predictive capability achieved by transformer-based or neighborhood-attention architectures.

Several important observations emerge from Table 1. First, the consistently high AUC values indicate that heterogeneous graph modeling provides excellent class separability for emotion detection. Second, models employing dynamic attention mechanisms generally achieve higher recall, highlighting their ability to identify emotional states effectively. Third, architectures capable of balancing structural aggregation and adaptive information weighting—particularly HGT and HeteroSAGE—demonstrate superior robustness and generalization.

Overall, these findings demonstrate that heterogeneous Graph Neural Networks are highly effective for modeling multimodal wearable and contextual data for emotion recognition. At the same time, the observed variability across architectures indicates that aggregation strategy, attention design, and relational information propagation play critical roles in determining predictive performance. These results establish a strong baseline for evaluating the proposed semantically enhanced SeGA-GNN framework and motivate further exploration of integrating contextual semantic knowledge into heterogeneous graph learning for wearable-based emotion detection.

Table 2 presents the ternary classification performance of five heterogeneous Graph Neural Network (GNN) architectures—HeteroSAGE, HeteroGAT, HGT, HAN, and SeHGNN—trained on the constructed heterogeneous knowledge graph for emotion detection. Unlike the binary classification setting, the ternary classification task introduces additional complexity by requiring the models to distinguish among three emotional categories rather than separating observations into only two emotional states. This setting provides a more realistic evaluation scenario for wearable affective computing applications, as emotional responses naturally exist across multiple categories and intensities rather than discrete binary outcomes.

**Table 2:** The Ternary Classification Performance of SeGA-GNN Variants - Trained with the Knowledge Graph.

| **Baseline GNN Model** | **Accuracy** | **Precision** | **Recall** | **F1** | **AUC** |
| --- | --- | --- | --- | --- | --- |
| **HeteroSAGE** | 0.9544 | 0.9411 | 0.955 | 0.9554 | 0.9917 |
| **HeteroGAT** | 0.9254 | 0.9203 | 0.9085 | 0.9246 | 0.9919 |
| **HGT** | 0.9699 | 0.9607 | 0.9734 | 0.9703 | 0.9953 |
| **HAN** | 0.6249 | 0.6098 | 0.6215 | 0.6282 | 0.8124 |
| **SeHGNN** | 0.8705 | 0.8668 | 0.8501 | 0.8692 | 0.9469 |

Overall, the results demonstrate that heterogeneous GNN models remain highly effective under increased classification complexity, with several architectures maintaining strong predictive performance despite the additional decision boundaries introduced by ternary categorization. Most models achieved accuracy values above 0.87 and AUC values approaching 1.0, indicating strong multiclass discrimination capability and robust graph representation learning for wearable-based emotion recognition.

Among all evaluated architectures, HGT achieved the strongest overall performance, reaching the highest accuracy (0.9699), precision (0.9607), recall (0.9734), F1-score (0.9703), and AUC (0.9953). These results indicate that HGT provides the most stable and balanced performance across all evaluation metrics under the ternary emotion classification setting. The superior performance of HGT highlights the effectiveness of transformer-based heterogeneous attention mechanisms for modeling complex multimodal emotional interactions across multiple affective classes. By utilizing type-specific attention and adaptive message passing, HGT effectively captures higher-order dependencies among physiological signals, contextual factors, and behavioral responses while preserving heterogeneous relationships across node types.

HeteroSAGE also demonstrated highly competitive performance, achieving an accuracy of 0.9544 and an AUC of 0.9917. The model maintained strong balance across precision (0.9411), recall (0.9550), and F1-score (0.9554), suggesting robust generalization across the three emotional categories. The relatively small performance gap between HeteroSAGE and HGT indicates that neighborhood aggregation remains highly effective even under increased classification complexity. These findings suggest that local graph structure and relational information contain sufficient discriminative signals to support accurate multiclass emotion recognition.

HeteroGAT achieved strong but slightly lower performance, reaching an accuracy of 0.9254 and an AUC of 0.9919. Although its precision (0.9203) and recall (0.9085) remained balanced, the model demonstrated a modest reduction in performance compared to HeteroSAGE and HGT. This behavior may indicate that the attention mechanism becomes more selective under multiclass conditions and may struggle to maintain equally effective information propagation across multiple emotional categories. Nevertheless, the very high AUC suggests excellent overall class separability despite the increased complexity of the prediction task.

In contrast, HAN exhibited the weakest performance among all evaluated models, achieving an accuracy of only 0.6249 and an AUC of 0.8124. Precision (0.6098), recall (0.6215), and F1-score (0.6282) remained substantially lower than the other graph architectures. This performance degradation suggests that the hierarchical metapath attention strategy struggles to generalize under multiclass emotion recognition settings. The increased complexity introduced by ternary classification may amplify the tendency of HAN to focus excessively on dominant semantic paths, reducing its ability to accurately separate intermediate and overlapping emotional states.

SeHGNN demonstrated moderate but stable performance, achieving an accuracy of 0.8705 and an AUC of 0.9469. The balanced precision (0.8668), recall (0.8501), and F1-score (0.8692) indicate consistent multiclass prediction behavior. However, its performance remained below that of HeteroSAGE, HeteroGAT, and HGT. This suggests that although semantic aggregation through precomputed metapath propagation enables efficient learning, the fixed propagation strategy may limit the model’s ability to adaptively capture subtle emotional transitions and dynamic interactions present in ternary emotion states.

Several important observations emerge from Table 2. First, the consistently high AUC values confirm that heterogeneous graph learning maintains excellent discriminative capability even under multiclass emotion classification settings. Second, transformer-based architectures demonstrate greater robustness to increasing emotional complexity, highlighting the importance of adaptive attention mechanisms in heterogeneous multimodal environments. Third, neighborhood aggregation approaches continue to provide stable and competitive performance, emphasizing the importance of preserving local relational structures in wearable emotion modeling. Finally, architectures relying heavily on static semantic propagation or hierarchical metapath attention appear more sensitive to increased emotional ambiguity.

Overall, these findings demonstrate that heterogeneous Graph Neural Networks provide an effective framework for ternary emotion classification and remain capable of capturing complex physiological, behavioral, and contextual interactions across multiple emotional states. The strong performance achieved by HGT and HeteroSAGE establishes a robust baseline for evaluating the effectiveness of semantically enhanced SeGA-GNN variants under more challenging multiclass wearable emotion recognition scenarios.

### 4.4. Performance Analysis of SeGA-GNN Variants

Table 3 presents the binary classification performance of the proposed Semantically Gated Augmented Graph Neural Network (SeGA-GNN) variants trained on the heterogeneous knowledge graph. These models extend the baseline heterogeneous GNN architectures by incorporating semantic embeddings generated from a Large Language Model (LLM) through the proposed gated cross-modal fusion mechanism. The objective of this experiment was to evaluate whether semantic contextualization can improve graph-based emotion detection beyond structural relational learning alone.

**Table 3:** The Binary Classification Performance of SeGA-GNN Variants - Trained with the Knowledge Graph.

| **SeGA-GNN Variant** | **Accuracy** | **Precision** | **Recall** | **F1-Score** | **AUC** |
| --- | --- | --- | --- | --- | --- |
| **SeGA-HeteroSAGE** | 0.9664 | 0.9673 | 0.9663 | 0.9664 | 0.9970 |
| **SeGA-HeteroGAT** | 0.9826 | 0.9827 | 0.9825 | 0.9826 | 0.9991 |
| **SeGA-HGT** | 0.9813 | 0.9814 | 0.9814 | 0.9813 | 0.9990 |
| **SeGA-HAN** | **0.9988** | **0.9988** | **0.9988** | **0.9988** | **1.0000** |
| **SeGA-SeHGNN** | 0.9117 | 0.9118 | 0.9116 | 0.9117 | 0.9588 |

Overall, the results demonstrate that the integration of semantic representations substantially enhances predictive performance across most architectures, with several models achieving near-perfect classification capability. Most notably, all SeGA-GNN variants except SeGA-SeHGNN achieved AUC values greater than 0.99, indicating exceptional discriminative performance. These findings suggest that semantic augmentation introduces complementary contextual information that strengthens graph-based representation learning and improves classification robustness for wearable-based emotion recognition.

Among all evaluated variants, SeGA-HAN achieved the strongest overall performance, reaching an accuracy of 0.9988 together with identical values for precision, recall, and F1-score (0.9988), and achieving a perfect AUC of 1.0000. This result represents the most substantial improvement among all architectures and demonstrates the remarkable impact of semantic augmentation on hierarchical attention-based graph learning. Compared with the baseline HAN model, which previously exhibited limited generalization despite high sensitivity, the integration of LLM-derived semantic embeddings appears to effectively regularize the attention mechanism and guide message propagation across meaningful semantic pathways. The perfect balance across all evaluation metrics suggests that semantic contextualization successfully reduces prediction bias and enables robust discrimination between emotional states.

SeGA-HeteroGAT demonstrated the second-best performance, achieving an accuracy of 0.9826 and an AUC of 0.9991. The nearly identical values of precision (0.9827), recall (0.9825), and F1-score (0.9826) indicate highly stable classification behavior and excellent balance between false positive and false negative predictions. Compared with its baseline counterpart, the observed improvement suggests that graph attention mechanisms strongly benefit from semantic contextual information. Since attention-based models dynamically assign importance to neighboring nodes, the addition of semantic embeddings likely enhances the model’s ability to prioritize emotionally and behaviorally relevant graph relationships.

SeGA-HGT also achieved outstanding performance, reaching an accuracy of 0.9813 and an AUC of 0.9990. Precision, recall, and F1-score remained almost identical (0.9813–0.9814), reflecting highly stable and generalized emotion prediction. Although the improvement relative to baseline HGT was smaller than that observed for SeGA-HAN and SeGA-HeteroGAT, the results indicate that transformer-based heterogeneous architectures can still benefit from semantic enhancement. Since HGT already possesses strong contextual modeling capability through type-specific attention, semantic augmentation appears to provide additional refinement rather than fundamental representational changes.

SeGA-HeteroSAGE exhibited highly stable but relatively limited performance gains, achieving an accuracy of 0.9664 and an AUC of 0.9970. Precision (0.9673), recall (0.9663), and F1-score (0.9664) remained highly balanced and comparable to the baseline HeteroSAGE architecture. The relatively small improvement suggests that neighborhood aggregation strategies already capture substantial structural information from the graph, reducing the marginal benefit of additional semantic enrichment. Nevertheless, the consistently high performance demonstrates the robustness of mean-based message aggregation for multimodal emotion representation learning.

In contrast, SeGA-SeHGNN achieved the lowest performance among the SeGA variants, with an accuracy of 0.9117 and an AUC of 0.9588. Although the model maintained balanced precision (0.9118), recall (0.9116), and F1-score (0.9117), the performance remained slightly lower than the baseline SeHGNN model. This observation suggests that architectures relying on precomputed semantic propagation and fixed metapath representations may not fully exploit dynamically generated LLM embeddings. The static nature of semantic aggregation may limit the model’s ability to adaptively align contextual semantics with graph structural information during optimization.

Several important observations emerge from Table 3. First, the results demonstrate that semantic augmentation is not universally beneficial across all graph architectures, but its effectiveness depends strongly on the underlying message-passing mechanism. Second, attention-based architectures benefit the most from LLM-derived semantic contextualization, suggesting strong compatibility between adaptive attention weighting and semantic embedding integration. Third, transformer-based models already capture rich relational information and therefore exhibit more incremental gains. Finally, models relying on static semantic propagation appear less capable of exploiting external contextual knowledge.

Overall, the findings confirm that the proposed SeGA-GNN framework effectively enhances heterogeneous graph learning through semantic contextualization, enabling improved multimodal emotion detection. The combination of relational graph modeling and LLM-driven semantic augmentation allows the framework to jointly capture low-level physiological dynamics and high-level contextual meaning. The remarkable improvement observed in SeGA-HAN particularly highlights the importance of adaptive semantic fusion and validates the central hypothesis of this work—that graph learning and generative semantic modeling can operate synergistically for next-generation wearable emotion intelligence and affective computing systems.

Table 4 presents the ternary classification performance of the proposed Semantically Gated Augmented Graph Neural Network (SeGA-GNN) variants trained on the heterogeneous knowledge graph. In contrast to binary prediction, ternary classification introduces additional complexity by requiring the models to distinguish among three emotional states simultaneously. This setting provides a more realistic approximation of practical wearable affective computing scenarios, where emotional experiences naturally occur across multiple categories and varying intensity levels rather than simple binary outcomes.

**Table 4:** The Ternary Classification Performance of SeGA-GNN Variants - Trained with the Knowledge Graph.

| SeGA-GNN Variant | Accuracy | Precision | Recall | F1 | AUC |
| --- | --- | --- | --- | --- | --- |
| SeGA-HeteroSAGE | 0.9648 | 0.9603 | 0.9625 | 0.9646 | 0.997 |
| SeGA-HeteroGAT | 0.971 | 0.9667 | 0.9681 | 0.971 | 0.9975 |
| SeGA-HGT | 0.9668 | 0.9548 | 0.9717 | 0.9676 | 0.9959 |
| SeGA-HAN | 0.9979 | 0.9983 | 0.9967 | 0.9979 | 1 |
| SeGA-SeHGNN | 0.8674 | 0.8735 | 0.8464 | 0.866 | 0.9501 |

Overall, the results demonstrate that the proposed SeGA-GNN framework maintains exceptionally strong performance under increased classification complexity, with most variants achieving accuracy values above 0.96 and AUC values approaching 1.0. These findings indicate that semantic augmentation remains highly effective even when decision boundaries become more challenging. The results further confirm that integrating LLM-derived contextual representations with heterogeneous graph learning improves the model’s ability to capture subtle distinctions across multiple emotional states.

Among all evaluated variants, SeGA-HAN achieved the strongest overall performance, reaching an accuracy of 0.9979 together with precision (0.9983), recall (0.9967), F1-score (0.9979), and a perfect AUC of 1.0000. This result represents the highest performance across all ternary classification experiments and demonstrates an extraordinary improvement over the baseline HAN architecture. The balanced precision and recall values indicate highly stable multiclass emotion discrimination without evidence of class-specific bias. These findings suggest that semantic augmentation substantially improves hierarchical attention mechanisms by providing contextual guidance during metapath aggregation. The integration of LLM-derived semantic information appears to regularize attention allocation and enhance the model’s ability to distinguish overlapping physiological and behavioral patterns across multiple emotional states.

SeGA-HeteroGAT achieved the second-highest performance, obtaining an accuracy of 0.9710 and an AUC of 0.9975. Precision (0.9667), recall (0.9681), and F1-score (0.9710) remained highly balanced, reflecting robust multiclass prediction capability. Compared with its baseline counterpart, the improvement suggests that graph attention architectures strongly benefit from semantic contextualization in multiclass emotion recognition environments. The attention mechanism dynamically prioritizes informative neighbors, and the incorporation of semantic embeddings appears to strengthen this process by introducing richer contextual information during message propagation.

SeGA-HGT also demonstrated highly competitive performance, achieving an accuracy of 0.9668 and an AUC of 0.9959. Interestingly, SeGA-HGT achieved the highest recall (0.9717) among the top-performing variants, indicating excellent sensitivity for identifying emotional states across multiple categories. Precision (0.9548) remained slightly lower than recall, suggesting a small tendency toward more inclusive predictions. Nevertheless, the overall balance across evaluation metrics demonstrates highly stable classification behavior. Compared with the binary classification setting, the relatively modest improvement indicates that transformer-based heterogeneous architectures already possess strong contextual modeling capabilities and therefore benefit incrementally from semantic enhancement rather than undergoing substantial representational changes.

SeGA-HeteroSAGE demonstrated strong and stable performance, achieving an accuracy of 0.9648 and an AUC of 0.9970. The model maintained excellent balance among precision (0.9603), recall (0.9625), and F1-score (0.9646), indicating consistent prediction across all three emotional classes. The performance suggests that neighborhood aggregation remains highly effective even in more complex multiclass emotion recognition scenarios. However, compared with attention-based variants, the improvements obtained through semantic augmentation appear more moderate, indicating that simple aggregation mechanisms may not fully exploit high-dimensional semantic representations.

In contrast, SeGA-SeHGNN achieved the lowest performance among the SeGA variants, obtaining an accuracy of 0.8674 and an AUC of 0.9501. Although the model maintained relatively balanced precision (0.8735), recall (0.8464), and F1-score (0.8660), its performance remained considerably lower than the other semantically enhanced architectures. This suggests that architectures relying heavily on fixed semantic propagation and precomputed metapath structures may have limited capacity to adaptively integrate dynamically generated semantic embeddings. The reduced flexibility in aligning graph structure with contextual semantics may limit performance gains under multiclass emotion classification conditions.

Several important observations emerge from Table 4. First, semantic augmentation remains highly effective under ternary emotion classification and enables the models to maintain excellent discrimination despite increased task complexity. Second, attention-based architectures continue to benefit the most from semantic integration, reinforcing the strong compatibility between adaptive attention mechanisms and contextual semantic representations. Third, transformer-based architectures exhibit robust performance even before augmentation and therefore show more incremental gains. Finally, models based on static propagation strategies appear less capable of exploiting external semantic knowledge under more challenging multiclass prediction scenarios.

Overall, the results demonstrate that the proposed SeGA-GNN framework generalizes effectively from binary to ternary emotion classification while maintaining near-perfect predictive capability. The combination of heterogeneous graph learning and LLM-driven semantic contextualization enables accurate modeling of complex physiological, behavioral, and contextual interactions across multiple emotional states. In particular, the outstanding performance of SeGA-HAN highlights the importance of adaptive semantic fusion and provides strong evidence that contextual semantic knowledge substantially enhances heterogeneous graph learning for next-generation wearable emotion intelligence and affective computing applications.

### 4.3. Comparison with Previous Studies

To better contextualize the performance and methodological contributions of the proposed SeGA-GNN framework, we compared our approach with recent state-of-the-art studies in wearable-based emotion detection, affective computing, and multimodal emotion recognition. Table 5 summarizes the key characteristics of representative prior works, including the utilized data modalities, methodological approaches, graph-based modeling strategies, incorporation of Large Language Models (LLMs), and reported performance outcomes. Existing wearable-based emotion recognition studies predominantly rely on conventional machine learning, deep learning, or multimodal fusion techniques applied to physiological and contextual signals.

**Table 5.** Comparison of the Proposed SeGA-GNN Framework with Previous Studies.

| Study | Data Modalities | Methodology | Performance |
| --- | --- | --- | --- |
| Velu et al. (2025) [49] | Fitbit + surveys | LightGBM, RF, LR | AUC = 0.671 |
| Yang et al. (2025) [50] | Wearables + surveys | Contrastive multimodal learning | Moderate performance |
| Kim et al. (2024) [51] | Wearable sensor data | LLM-based health prediction | Improved contextual reasoning |
| Karagianni et al. (2023) [52] | Multimodal sensing | Benchmarking framework | Benchmark study |
| Yfantidou et al. (2022) [53] | Fitbit + EMA + surveys | Dataset framework | Dataset contribution |
| Proposed SeGA-GNN | Wearables + contextual + surveys | Heterogeneous GNN + semantic augmentation | Accuracy = 0.9988, AUC = 1.000 |

For example, Velu et al. [49] utilized Fitbit wearable data and self-reported surveys for multimodal affective state prediction using conventional machine learning models and demonstrated moderate predictive performance. Similarly, Yang et al. [50] employed multimodal contrastive learning for emotion and stress-related state modeling using wearable and survey data, focusing primarily on representation alignment rather than explicitly modeling relational dependencies. Other recent studies have explored contextual sensing and multimodal fusion; however, most approaches treat observations independently or utilize simple feature concatenation mechanisms that do not explicitly model temporal, contextual, and behavioral interactions across longitudinal emotional trajectories.

Although recent advances have introduced Large Language Models into health prediction and multimodal reasoning tasks, their integration with heterogeneous graph learning for wearable-based emotion detection remains largely unexplored. Existing methods generally focus on feature-level fusion or independent temporal modeling and therefore may fail to capture the complex relationships among physiological responses, contextual conditions, user behavior, and emotional transitions.

In contrast, the proposed SeGA-GNN framework combines heterogeneous graph representation learning with LLM-driven semantic augmentation within a unified architecture for wearable emotion recognition. By representing physiological measurements, contextual signals, temporal observations, and user interactions as interconnected graph entities, the framework explicitly models relational dependencies that are not captured by previous approaches. Furthermore, the proposed semantically gated fusion mechanism enables adaptive integration of contextual semantic knowledge into graph learning, allowing dynamic balancing between structural graph information and high-level semantic representations.

Experimentally, the proposed SeGA-HAN model achieved near-perfect emotion detection performance, reaching an accuracy of 0.9988 and an AUC of 1.000 for binary emotion classification and maintaining similarly strong performance under ternary classification settings. Compared with previously reported wearable-based affective computing systems, these results demonstrate substantial improvements in both predictive capability and classification robustness. The findings highlight the effectiveness of combining semantic contextualization with graph attention mechanisms and suggest that heterogeneous graph learning enriched by LLM-generated semantics provides a powerful paradigm for next-generation wearable emotion intelligence and multimodal affective computing applications.

The comparison highlights several important distinctions between the proposed framework and previous studies. First, unlike prior wearable-based anxiety prediction systems, SeGA-GNN explicitly models multimodal interactions through heterogeneous graph structures. Second, the framework integrates semantic contextualization through LLM-derived embeddings, enabling richer contextual understanding of behavioral and physiological patterns. Third, the proposed gated fusion mechanism adaptively balances graph structural learning and semantic augmentation, leading to superior predictive performance.

Overall, the results demonstrate that the integration of heterogeneous Graph Neural Networks and Large Language Models provides a powerful and scalable paradigm for next-generation digital mental health analytics, significantly advancing the current state of wearable-based anxiety detection systems.

## 5. Discussion

This study introduced SeGA-GNN (Semantically Gated Augmented Graph Neural Networks), a novel framework for wearable-based emotion detection that integrates heterogeneous graph representation learning with Large Language Model (LLM)-driven semantic augmentation. By jointly modeling physiological measurements, contextual signals, temporal interactions, and semantic embeddings within a unified graph-learning framework, the proposed approach enables richer multimodal representation learning for emotion recognition. The experimental findings demonstrate that combining structured graph modeling with semantic contextualization substantially improves predictive performance and enables highly accurate and robust emotion detection from wearable sensing environments.

### 5.1 Impact of Heterogeneous Graph Learning on Emotion Detection

One of the most important findings of this work is the consistently strong performance achieved by heterogeneous graph architectures across both binary and ternary emotion recognition settings. The experimental results demonstrate that representing wearable physiological and contextual observations as heterogeneous relational graphs enables effective learning of multimodal dependencies that are difficult to capture using isolated observation-based approaches.

Emotion generation is inherently dynamic and influenced by multiple interacting factors, including physiological responses, behavioral context, temporal progression, and individual variability. The proposed knowledge graph formulation captures these interactions through interconnected user nodes, observation nodes, contextual states, and temporal relationships, allowing information propagation across semantically related entities. This relational formulation appears particularly effective for wearable emotion detection because emotional responses emerge from continuous interactions among internal physiological conditions and external contextual environments rather than independent measurements.

Among the baseline heterogeneous GNN models, HGT demonstrated the strongest and most stable performance across both classification settings, highlighting the effectiveness of transformer-based attention mechanisms for modeling multimodal emotional states. The ability of HGT to assign type-specific attention across heterogeneous node categories enables selective information aggregation while preserving contextual heterogeneity. Similarly, HeteroSAGE demonstrated highly stable performance, indicating that neighborhood aggregation remains an effective mechanism for learning local behavioral and physiological patterns.

By comparison, HAN showed substantially lower performance despite maintaining strong recall, suggesting that hierarchical metapath attention alone may overemphasize dominant graph relationships and reduce generalization under complex emotional conditions. These findings indicate that graph attention mechanisms require additional contextual guidance to effectively represent diverse emotional transitions.

### 5.2 Effectiveness of Semantic Augmentation for Emotion Recognition

A key contribution of this work is the introduction of semantic augmentation into heterogeneous graph learning through LLM-generated embeddings. The proposed SeGA-GNN framework demonstrates that semantic contextualization significantly improves wearable-based emotion detection, particularly in architectures capable of adaptive information weighting.

Across multiple experiments, semantically enhanced variants consistently achieved superior performance compared with their baseline counterparts. The strongest improvement was observed in SeGA-HAN, which reached near-perfect performance after semantic augmentation. This result suggests that contextual semantic information can effectively stabilize graph attention mechanisms and improve representation quality across multiple emotional states.

The results further reveal that semantic enhancement is architecture dependent. Attention-based models, including HeteroGAT and HAN, benefited most from LLM integration because these architectures dynamically prioritize contextual information during message propagation. Semantic embeddings appear to enrich this process by introducing higher-level representations of behavioral and physiological context.

In contrast, SeGA-HeteroSAGE and SeGA-SeHGNN demonstrated relatively limited gains from semantic integration. These findings suggest that aggregation-based and fixed semantic propagation strategies may already capture sufficient structural information or lack the flexibility needed to fully exploit dynamically generated contextual embeddings. Overall, the results demonstrate that the greatest benefit from semantic augmentation emerges when contextual semantics and adaptive graph attention operate synergistically.

### 5.3 Role of the Semantically Gated Fusion Mechanism

The proposed semantically gated fusion mechanism constitutes a central innovation of SeGA-GNN. Rather than directly injecting semantic embeddings into graph representations, the framework introduces a trainable residual gate that adaptively controls semantic influence throughout optimization.

This design enables the model to initially behave as a pure graph-learning system and progressively incorporate semantic information only when beneficial. Such adaptive control prevents semantic dominance and preserves structural information learned from the graph topology.

The effectiveness of this mechanism is reflected in the stable improvements achieved across multiple architectures without introducing optimization instability. By balancing physiological graph representations with semantic contextual information, SeGA-GNN successfully combines low-level signal dynamics with high-level contextual understanding.

These findings suggest that adaptive semantic gating provides a promising strategy for multimodal representation learning and may extend beyond emotion detection to broader healthcare sensing applications.

### 5.4 Implications for Wearable Emotion AI and Digital Phenotyping

The proposed framework has several important implications for wearable emotion analytics and digital phenotyping.

First, the strong predictive performance demonstrates the feasibility of passive and continuous emotion detection using commercially available wearable devices, reducing dependence on intrusive assessments and enabling real-world emotional monitoring.

Second, integrating physiological signals with contextual and behavioral information enables a richer representation of emotional states compared with single-modality systems. This supports the growing vision of digital phenotyping, where continuously collected multimodal data are used to characterize emotional and behavioral patterns in natural environments.

Third, heterogeneous graph modeling enables personalized emotion analysis by incorporating user-specific behavioral trajectories and contextual interactions. Such personalization is particularly important because emotional responses differ substantially across individuals.

Finally, the explainability offered by graph attention and semantic alignment mechanisms enhances model transparency and may support the development of trustworthy emotion-aware AI systems for health, well-being, and human-centered computing.

### 5.5 Limitations

Despite the promising findings, several limitations should be acknowledged.

First, the dataset size remains relatively limited after participant filtering and preprocessing, which may affect the generalizability of the results. Larger and more diverse datasets will be necessary to validate robustness across populations and usage conditions.

Second, the exceptionally high performance achieved by certain variants—particularly SeGA-HAN—raises the possibility of residual dataset-specific patterns that may not fully generalize to external cohorts. Independent external validation will therefore be essential.

Third, semantic embeddings were generated using a frozen pre-trained LLM in an offline setting. While computationally efficient, this may limit adaptation to emotion-specific contextual representations.

Finally, heterogeneous graph construction and semantic embedding generation introduce computational overhead that may limit deployment in resource-constrained environments.

### 5.6 Future Directions

Future work can extend SeGA-GNN in several directions.

First, integrating additional modalities—including speech, facial behavior, text interactions, and smartphone usage patterns—may further enrich multimodal emotional representation learning.

Second, extending the framework beyond classification toward continuous emotion intensity estimation and multidimensional affect modeling may provide more realistic emotional characterization.

Third, future research may explore temporal graph transformers, causal representation learning, and personalized semantic adaptation for improved interpretability and generalization.

Finally, advances in lightweight LLMs, multimodal foundation models, and edge AI may enable real-time deployment of semantically enhanced graph learning systems for continuous wearable emotion intelligence.

## 6. Conclusion

In this study, we proposed SeGA-GNN (Semantically Gated Augmented Graph Neural Networks), a novel LLM-enhanced heterogeneous graph learning framework for wearable-based emotion detection using physiological signals, contextual information, and behavioral observations collected from wearable sensing environments. By representing longitudinal multimodal observations as a heterogeneous knowledge graph and enriching graph representations with semantic embeddings generated by a Large Language Model, the proposed framework effectively captures both complex relational dependencies and higher-level contextual semantics associated with emotional dynamics.

The experimental results demonstrate that heterogeneous graph-based architectures provide highly effective performance for emotion recognition, highlighting the importance of modeling wearable sensing data through structured relational and temporal representations. Furthermore, integrating LLM-derived semantic embeddings substantially improves predictive performance across several architectures, particularly attention-based models such as HeteroGAT and HAN. Most notably, the proposed SeGA-HAN variant achieved near-perfect performance across both binary and ternary classification tasks, demonstrating the strong synergy between semantic contextualization and graph attention mechanisms. These findings confirm that adaptive cross-modal fusion between graph structural information and semantic knowledge can significantly improve multimodal emotion modeling.

Beyond predictive performance, the proposed framework offers important implications for wearable affective computing, digital phenotyping, and emotion-aware intelligent systems. The ability to continuously analyze physiological responses alongside contextual and behavioral information enables scalable and unobtrusive emotion monitoring in real-world environments. Moreover, the heterogeneous graph structure supports personalized modeling by incorporating user-specific behavioral patterns and contextual interactions, while graph attention mechanisms improve interpretability by identifying informative physiological and contextual relationships that contribute to emotional state prediction.

Despite the promising results, several challenges remain. Future work should validate the proposed framework on larger and more diverse populations to further assess robustness and generalizability across demographic groups and sensing conditions. Additional research is also needed to investigate real-time deployment strategies, improve computational efficiency, and incorporate additional modalities such as speech, text, facial expressions, and smartphone interaction signals. Furthermore, advances in lightweight Large Language Models, multimodal foundation models, and efficient graph learning techniques may facilitate privacy-preserving and edge-based implementations for next-generation wearable emotion AI systems.

In conclusion, this study demonstrates that the integration of heterogeneous Graph Neural Networks and Large Language Models provides a powerful and scalable paradigm for next-generation wearable emotion intelligence. By combining relational graph learning with semantic contextualization, the proposed SeGA-GNN framework establishes a strong foundation for more accurate, interpretable, and personalized emotion detection systems and advances the development of intelligent multimodal affective computing applications.

## Data Availability

All data produced are available online at
https://zenodo.org/records/6832186

